# Ischemic stroke after COVID-19 bivalent vaccine administration in patients aged 65 years and older: analysis of nation-wide patient electronic health records in the United States

**DOI:** 10.1101/2023.02.11.23285801

**Authors:** Maria P. Gorenflo, Pamela B. Davis, David C. Kaelber, Rong Xu

## Abstract

**Importance:** The Centers for Disease Control and Prevention (CDC) announced in January 2023 that they were investigating a potential connection between administration of the Pfizer novel coronavirus disease-2019 (COVID-19) bivalent vaccine booster and ischemic stroke (IS).

**Objective:** To explore the relationship between Pfizer bivalent booster administration and IS in older patients in the United States and compare it to other COVID-19 vaccines.

**Design:** A retrospective cohort study was conducted to compare hazard of IS among patients aged 65 years or over who received the Pfizer bivalent, Moderna bivalent, or Pfizer/Moderna monovalent COVID-19 booster vaccine 1-21 and 22-42 days after vaccination.

**Setting:** Patient data were collected from TriNetX, a cloud-based analytics platform that includes electronic health record data from over 90 million unique patients in the United States.

**Participants:** Patients in the United States aged 65 years or over at the time of administration of a Pfizer bivalent (n = 43,216), Moderna bivalent (n = 4,267), or Pfizer/Moderna monovalent (n = 100,583) booster were included for analysis. Cohorts were propensity-score matched by demographic factors and risk factors for IS and severe COVID-19.

**Exposures:** Pfizer bivalent, Moderna bivalent, or Pfizer/Moderna monovalent COVID-19 booster administration.

**Main outcomes:** The hazard ratio (HR) and 95% confidence interval (CI) for IS in the cohorts at 1-21 and 22-42 days after administration.

**Results:** After matching, the Pfizer bivalent cohort included 4,267 patients, with an average age of 73.7 years (44.43% male, 76.59% white). The Moderna bivalent cohort included 4,267 patients, with an average age of 74.0 years (44.08% male, 77.39% white). There was no significant difference in the hazard of IS encounters between the Pfizer bivalent versus Moderna bivalent cohorts at 1-21- or 22-42-days post-administration: HR = 0.59 (0.31, 1.11), 0.73 (0.33, 1.60). The hazard for IS was lower in the Pfizer bivalent cohort than in the Pfizer/Moderna monovalent cohort at both timepoints: HR = 0.24 (0.19, 0.29), 0.25 (0.20, 0.31).

**Conclusions and relevance:** Older adults administered the Pfizer bivalent booster had similar hazard for IS encounters compared to those administered the Moderna bivalent booster vaccine, but lower hazard than those administered the Pfizer/Moderna monovalent boosters.

**Key Points:** *Question:* What is the comparative hazard of ischemic stroke in American patients ages 65 years and over after administration of the Pfizer bivalent, Moderna bivalent, or Pfizer/Moderna monovalent COVID-19 booster vaccine?

*Findings:* A retrospective cohort study was conducted. There was no significant difference in the hazard of ischemic stroke encounters between the Pfizer bivalent versus Moderna bivalent cohorts, but lower hazard for the Pfizer bivalent than the monovalent boosters at 1-21 or 22-42 days post-administration.

*Meaning:* There is no evidence from these results that the Pfizer bivalent booster is associated with increased hazard for ischemic stroke.

## 1. Introduction

On January 13, 2023, the Centers for Disease Control and Prevention (CDC) announced that its Vaccine Safety Datalink met the threshold to investigate the risk of ischemic stroke (IS) within three weeks of administration of the Pfizer/BioNTech novel coronavirus disease-2019 (COVID-19) bivalent vaccine (Pfizer bivalent booster)^1^. No such concern was raised in the CDC’s statement for the Moderna COVID-19 bivalent vaccine (Moderna bivalent booster). The earlier monovalent Moderna and Pfizer vaccines display no increased risk for IS in the general American population^2^; however, COVID-19 infection itself appears to be a risk factor for IS in patients ages 65 years and over^3^. This CDC announcement was therefore unanticipated, since then both the Food and Drug Administration and European Medicines Agency have reported no increased IS risk for the Pfizer bivalent vaccine in their respective databases^4,5^. In response to the inconsistent findings among these major regulatory bodies and the wide use of COVID-19 bivalent vaccines in older adults, we set out to gauge the hazard of IS in patients ages 65 years and over administered the Pfizer bivalent booster in a nationwide, frequently updated platform of electronic health record (EHR) data on over 90 million unique patients in the United States.

## 2. Methods

### Data collection, study population, variables, and outcomes

We used the TriNetX platform to access aggregated, de-identified EHRs of 90 million patients from 56 health care organizations across all 50 American states, covering diverse geographic, age, race, and ethnic groups (United States Collaborative Network)^5^ (eMethod). The MetroHealth System, Cleveland Ohio, Institutional Review Board has determined that research using TriNetX for retrospective cohort studies is not Human Subject Research and is therefore exempt from review. We have previously used the TriNetX platform to study risk factors and outcomes of COVID-19 infection and vaccination^6–8^.

There were two components of this study: (1) a comparison of IS encounters in patients aged 65 years and over before and after Pfizer bivalent vaccine administration and (2) comparison of the hazards of IS after administration of the Pfizer bivalent versus Moderna bivalent boosters, and Pfizer bivalent versus Pfizer/Moderna monovalent boosters. The exposure of interest was vaccination by either the Pfizer bivalent booster (“Pfizer bivalent” cohort), Moderna bivalent booster (“Moderna bivalent” cohort), or Pfizer/Moderna monovalent booster (“monovalent cohort”) prior to November 25, 2022 to ensure sufficient time for follow-up at 21 and 42 days. Cohorts were matched by demographics (age, sex, race, ethnicity); COVID-19 infection; diagnostic risk factors for IS and severe COVID-19 infection (hypertension, overweight, obesity, and type II diabetes mellitus); and risk factors for IS alone (drug, alcohol, and nicotine use; low socioeconomic status; dyslipidemia, ischemic heart disease, atrial fibrillation, cerebral vascular disease, and previous IS)^9,10^ (eTable 1 in eMethod). The outcome of interest was any encounter for IS in TriNetX at either 1-21 days or 22-42 days after booster administration.

**Table 1:** Patient characteristics in the Pfizer and Moderna bivalent cohorts before and after propensity-score matching.

| Covariate | Before Matching, No. (%) |  |  | After Matching, No. (%) |  |  |
| --- | --- | --- | --- | --- | --- | --- |
|  | Pfizer bivalent cohort (n = 43,216) | Moderna bivalent cohort (n = 4,267) | SMD<sup>a</sup> | Pfizer bivalent cohort (n = 4,267) | Moderna bivalent cohort (n = 4,267) | SMD<sup>a</sup> |
| **Demographics** |  |  |  |  |  |  |
| Age at booster (years)<sup>b</sup> | 73.6 (6.05) | 74.0 (6.42) | 0.08 | 73.7 (6.36) | 74.0 (6.42) | 0.05 |
| Sex |  |  |  |  |  |  |
| Male | 19,726 (45.65) | 1,881 (44.07) | 0.03 | 1,896 (44.43) | 1,881 (44.08) | <0.01 |
| Female | 23,483 (54.34) | 2,387 (55.93) | 0.03 | 2,371 (55.57) | 2,386 (55.92) | <0.01 |
| Race |  |  |  |  |  |  |
| White | 35,382 (81.87) | 3,302 (77.37) | 0.11 | 3,268 (76.59) | 3,302 (77.39) | 0.02 |
| Black or African American | 3,977 (9.20) | 734 (17.20) | 0.24 | 765 (17.93) | 733 (17.18) | 0.02 |
| Asian | 587 (1.36) | 86 (2.02) | 0.05 | 83 (1.95) | 86 (2.02) | <0.01 |
| Unknown | 3,074 (7.11) | 138 (3.23) | 0.18 | 128 (3.00) | 138 (3.23) | 0.01 |
| Ethnicity |  |  |  |  |  |  |
| Hispanic or Latino | 3,525 (8.16) | 285 (6.68) | 0.06 | 302 (7.08) | 285 (6.68) | 0.02 |
| Not Hispanic or Latino | 37,827 (87.53) | 3,844 (90.07) | 0.08 | 3,827 (89.69) | 3,844 (90.09) | 0.01 |
| Unknown | 1,864 (4.31) | 139 (3.26) | 0.06 | 138 (3.23) | 138 (3.23) | <0.01 |
| **Diagnoses** |  |  |  |  |  |  |
| Type II diabetes mellitus | 14,862 (34.39) | 2,485 (58.22) | 0.49 | 2,503 (58.66) | 2,484 (58.21) | <0.01 |
| Overweight and obesity | 13,305 (30.79) | 2,034 (47.66) | 0.35 | 2,058 (48.23) | 2,033 (47.65) | 0.01 |
| Disorders of lipoprotein metabolism and other lipidemias | 30,867 (71.43) | 3,372 (79.01) | 0.18 | 3,365 (78.86) | 3,372 (79.03) | <0.01 |
| Mental and behavioral disorders due to psychoactive substance use | 11,553 (26.73) | 2,454 (57.50) | 0.66 | 2,467 (57.82) | 2,453 (57.49) | <0.01 |
| Alcohol related disorders | 2,988 (6.91) | 532 (12.47) | 0.19 | 556 (13.03) | 532 (12.47) | 0.02 |
| Nicotine dependence | 7,918 (18.32) | 2,113 (49.51) | 0.70 | 2,108 (49.40) | 2,112 (49.50) | <0.01 |
| Essential hypertension | 29,555 (68.39) | 3,523 (82.55) | 0.33 | 3,557 (83.36) | 3,522 (82.54) | 0.02 |
| Ischemic heart disease | 13,104 (30.32) | 1,950 (45.69) | 0.32 | 1,914 (44.86) | 1,949 (45.68) | 0.02 |
| Atrial fibrillation | 5,902 (13.66) | 700 (16.40) | 0.08 | 712 (16.69) | 700 (16.41) | <0.01 |
| Cerebrovascular diseases | 9,605 (22.22) | 1,732 (40.58) | 0.40 | 1,673 (39.21) | 1,731 (40.57) | 0.03 |
| Cerebral infarction | 5,827 (13.48) | 1,450 (33.97) | 0.50 | 1,381 (32.37) | 1,449 (33.96) | 0.03 |
| Persons with potential health hazards related to socioeconomic and psychosocial circumstances | 7,714 (17.85) | 2,067 (48.43) | 0.69 | 2,064 (48.37) | 2,066 (48.42) | <0.01 |
| COVID-19 infection | 5,959 (13.79) | 548 (12.84) | 0.03 | 492 (11.53) | 548 (12.84) | 0.04 |

<sup>a</sup>Standard mean difference, a metric used to declare imbalance when greater than 0.1.
<sup>b</sup>Average age (standard deviation) is reported for this measurement.

### Statistical analysis

To compare the number of IS encounters in the Pfizer bivalent cohort at 1-21 days and 22-42 days before and after booster administration, Pearson’s chi-squared test was conducted using R, version 3.6.3. To compare the hazard of IS encounters between the Pfizer bivalent and Moderna bivalent cohorts, as well as the Pfizer bivalent and monovalent cohorts, the cohorts were propensity-score matched (1:1 matching by nearest neighbor greedy matching algorithm with a caliper of 0.25 standard deviations) for the variables enumerated above. Kaplan-Meier survival analysis was used to estimate the probability of IS encounter at 1-21 days or 22-42 days after booster administration. Cox’s proportional hazards model then compared cohorts with the proportional hazard assumption tested with the generalized Schoenfeld approach. Hazard ratios (HR) and 95% confidence intervals (CI) were generated to compare outcomes. A sub-analysis was conducted to compare new diagnosis of IS between the Pfizer bivalent cohort and the monovalent cohort, but not between the Pfizer bivalent cohort and Moderna bivalent cohort due to limited sample sizes. All analyses were conducted on January 28, 2023 within the TriNetX platform with significance set at p-value < 0.05 (two-sided).

## 3. Results

The study population of patients who were age 65 years or over at the time of booster administration on or before November 25, 2022 included 43,216 who received the Pfizer bivalent booster, 4,267 who received the Moderna bivalent booster, and 100,583 who received a Pfizer or Moderna monovalent booster. The characteristics of the Pfizer bivalent and Moderna bivalent cohorts before and after propensity-score matching are shown in Table 1. After matching, the Pfizer bivalent cohort was comprised of 4,267 patients, with an average age of 73.7 years (44.43% male, 76.59% white) (Table 1). The Moderna bivalent cohort was comprised of 4,267 patients, with an average age of 74.0 years (44.08% male, 77.39% white) (Table 1). Details on propensity-score matching between the Pfizer bivalent and monovalent cohorts are available in eTable 2 (eMethod).

Within the Pfizer bivalent cohort of 43,216 patients, there is no increase in IS encounters within the 1-21 days after versus before booster administration (127 versus 141 IS encounters, P = 0.43). However, there are fewer IS encounters in the 22-42 days after versus before booster administration (109 vs 189 IS encounters, P <0.001).

There was no significant difference in the hazard of IS encounters between the Pfizer bivalent and Moderna bivalent cohorts at 1-21- or 22-42-days post-administration: HR = 0.59 (0.31, 1.11), 0.73 (0.33, 1.60), respectively (Figure 1). Compared to the monovalent cohort, there is reduced hazard of IS encounters in the Pfizer bivalent cohort at both timepoints: HR = 0.24 (0.19, 0.29), 0.25 (0.20, 0.31), respectively (Figure 1). There was also reduced hazard of first-time IS encounters in the Pfizer bivalent cohort compared to the monovalent cohort at both time points, but this was only significant at 22-42 days post-administration: HR = 0.63 (0.35, 1.15), 0.32 (0.17, 0.60), respectively (Figure 1).

**Figure 1:**
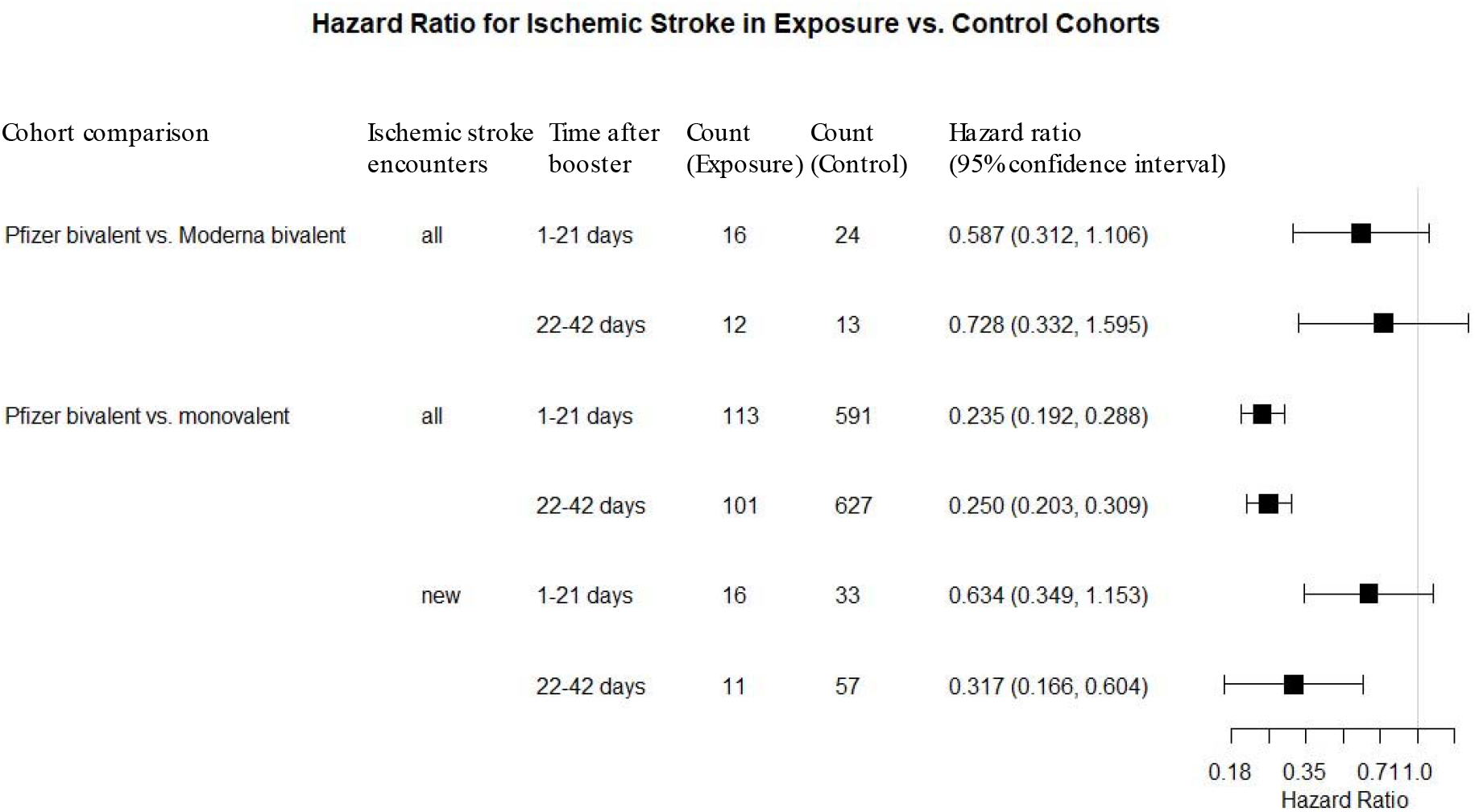
Hazard ratio for ischemic stroke between propensity-score matched cohorts 1-21 and 22-42 days after administration of Pfizer bivalent, Moderna bivalent, or Pfizer/Moderna monovalent COVID-19 vaccine.

## 4. Discussion

The hazard for IS encounters did not differ significantly between the matched Pfizer and Moderna bivalent cohorts at both 1-21 and 22-42 days after booster administration. We also observed a reduced hazard of IS encounters in the Pfizer bivalent versus monovalent cohort, perhaps due to bivalent boosters providing stronger protection against severe COVID-19 infection and hospitalization than monovalent vaccines^11,12^. Limitations of this study include the use of the TriNetX platform, which is not a random sampling of the entire United States population over the age of 65 years; therefore, the generalizability of the results is unclear. While the Pfizer and Moderna bivalent vaccines were approved in August 2022, their monovalent counterparts were approved earlier. Therefore, although patients in the Pfizer bivalent cohort and monovalent cohort were followed for the same length of time, the circulating SARS-Cov-2 virus variants that patients in these two cohorts encountered could be different. Nevertheless, our analysis provides no evidence that American patients ages 65 and over have an increased hazard of IS after Pfizer bivalent booster administration; patients and healthcare providers should not be dissuaded from receiving or administering this booster vaccine.

## Supporting information

eMethod

## Data Availability

All data produced in the present work are contained in the manuscript

## Contributors

RX conceived of, designed, and supervised the study and contributed to manuscript preparation. MPG designed and conducted the study and drafted the manuscript. PBD critically contributed to study design, result interpretation, and manuscript preparation. DCK provided access to the TriNetX platform and critically reviewed. All authors approved the final manuscript. We confirm the originality of content. MPG had full access to all the analysis in the study and takes responsibility for the integrity of the data and the accuracy of the data analysis.

## Acknowledgments

We acknowledge support from National Institute on Aging (grants nos. RF1AG076649), National Institute on Alcohol Abuse and Alcoholism (grant no. R01AA029831), and the Clinical and Translational Science Collaborative (CTSC) of Cleveland (grant no. 1UL1TR002548).

## Meeting Presentation

No

## References

1. CDC. COVID-19 Vaccination. Centers for Disease Control and Prevention. Published February 11, 2020. Accessed January 27, 2023. https://www.cdc.gov/coronavirus/2019-ncov/vaccines/safety/bivalent-boosters.html

2. Klein NP, Lewis N, Goddard K, et al. Surveillance for Adverse Events After COVID-19 mRNA Vaccination. JAMA. 2021;326(3):1390–1399. doi:10.1001/jama.2021.15072

3. Yang Q, Tong X, George MG, Chang A, Merritt RK. COVID-19 and Risk of Acute Ischemic Stroke Among Medicare Beneficiaries Aged 65 Years or Older. Neurology. 2022;98(3):e778. doi:10.1212/WNL.0000000000013184

4. Erman M. U.S. CDC still looking at potential stroke risk from Pfizer bivalent COVID shot. Reuters. https://www.reuters.com/business/healthcare-pharmaceuticals/us-cdc-still-looking-potential-stroke-risk-pfizer-bivalent-covid-shot-2023-01-26/. Published January 27, 2023. Accessed January 27, 2023.

5. Reuters. EU drug regulator has not seen signal of possible Pfizer COVID shot stroke link. Reuters. https://www.reuters.com/business/healthcare-pharmaceuticals/eu-drug-regulator-has-not-seen-signal-possible-pfizer-covid-shot-stroke-link-2023-01-18/. Published January 18, 2023. Accessed January 27, 2023.

6. Wang L, Davis PB, Kaelber DC, Volkow ND, Xu R. Comparison of mRNA-1273 and BNT162b2 Vaccines on Breakthrough COVID-19 Infections, Hospitalizations, and Death During the Delta-Predominant Period. JAMA. 2022;327(3):678–680. doi:10.1001/jama.2022.0210

7. Wang L, Berger NA, Kaelber DC, Davis PB, Volkow ND, Xu R. Incidence Rates and Clinical Outcomes of COVID-19 Infection With the Omicron and Delta Variants in Children Younger Than 5 Years in the US. JAMA Pediatr. 2022;176(3):811–813. doi:10.1001/jamapediatrics.2022.0945

8. Wang W, Kaelber DC, Xu R, Berger NA. Breakthrough COVID-19 Infections, Hospitalizations, and Mortality in Vaccinated Patients With Cancer in the US Between December 2020 and November 2021. JAMA Oncol. Published online April 8, 2022. doi:10.1001/jamaoncol.2022.1096

9. Boehme AK, Esenwa C, Elkind MSV. Stroke Risk Factors, Genetics, and Prevention. Circ Res. 2017;120(3):472–495. doi:10.1161/CIRCRESAHA.116.308398

10. Gao Y dong, Ding M, Dong X, et al. Risk factors for severe and critically ill COVID-19 patients: A review. Allergy. 2021;76(3):428–455. doi:10.1111/all.14657

11. Lin DY, Xu Y, Gu Y, et al. Effectiveness of Bivalent Boosters against Severe Omicron Infection. N Engl J Med. Published online January 25, 2023. doi:10.1056/NEJMc2215471

12. Collier A ris Y, Miller J, Hachmann NP, et al. Immunogenicity of BA.5 Bivalent mRNA Vaccine Boosters. N Engl J Med. Published online January 11, 2023. doi:10.1056/NEJMc2213948

