## Supplementary material for "Ischemic stroke after COVID-19 bivalent vaccine administration in patients aged 65 years and older: analysis of nation-wide patient electronic health records in the United States": eMethod

**Description of TriNetX database**

The data used in this study was collected on January 28, 2022 from the TriNetX “United States Collaborative Network” Network , which provided access to electronic heath records (diagnoses, procedures, medications, laboratory values, genomic information) from approximately 90 million patients from 56 healthcare organizations in the US across covering diverse geographic regions. TriNetX, LLC is compliant with the Health Insurance Portability and Accountability Act (HIPAA). Any data displayed on the TriNetX Platform in aggregate form, or any patient level data provided in a data set generated by the TriNetX Platform, only contains de-identified data as per the de-identification standard defined in Section §164.514(a) of the HIPAA Privacy Rule. MetroHealth System, Cleveland, Ohio, IRB has determined research using TriNetX, is not Human Subject Research and therefore exempt from IRB review.

TriNetX is a platform that de-identifies and aggregates electronic health record (EHR) data from contributing healthcare systems, most of which are large academic medical institutions with both inpatient and outpatient facilities at multiple locations, across 50 states in the US. TriNetX Analytics provides web-based and secure access to patient EHR data from hospitals, primary care, and specialty treatment providers, covering diverse geographic locations, age groups, racial and ethnic groups, income levels and insurance types including various commercial insurances, governmental insurance (Medicare and Medicaid), self-pay/uninsured, worker compensation insurance, military/VA insurance among others.

Self-reported race and ethnicity data in TriNetX comes from the underlying clinical EHR systems of the contributing healthcare systems. TriNetX maps race and ethnicity data from the contributing healthcare systems to the following categories: (1) Race: Asian, American Indian or Alaskan Native, Black or African American, Native Hawaiian or Other, White, Unknown race; and (2) Ethnicity: Hispanic or Latino, Not Hispanic or Latino, Unknown Ethnicity.

eTable 1: Codes for covariates, exposures, and outcomes used in TriNetX.

| Covariate | TriNetX Code |
|---|---|
| **Demographics** | **Code System Concept** |
| White | 2106-3 |
| Black or African American | 2054-5 |
| Asian | 2028-9 |
| Unknown Race | 2131-1 |

| | |
|---|---|
| Hispanic or Latino | 2135-2 |
| Not Hispanic or Latino | 2186-5 |
| **Diagnoses** | **International Classification of Diseases, 10th edition** |
| Type II diabetes mellitus | E11 |
| Overweight and obesity | E66 |
| Disorders of lipoprotein metabolism and other lipidemias | E78 |
| Mental and behavioral disorders due to psychoactive substance use | F10-F19 |
| Alcohol related disorders | F10 |
| Nicotine dependence | F17 |
| Essential hypertension | I10 |
| Ischemic heart disease | I20-I25 |
| Atrial fibrillation | I48 |
| Cerebrovascular diseases | I60-I69 |
| Cerebral infarction | I63 |
| Persons with potential health hazards related to socioeconomic and psychosocial circumstances | Z55-Z65 |
| COVID-19 | U07.1 |
| **Outcome** | **International Classification of Diseases, 10th edition** |
| Ischemic stroke | I63 |
| **Vaccination** | **Current Procedural Terminology** |
| Pfizer bivalent | 91312, 0124A |
| Moderna bivalent | 91313, 0134A |
| Pfizer monovalent | 91300 |
| Moderna monovalent | 91301 |

eTable 2: Patient characteristics in the Pfizer bivalent and monovalent cohorts before and after propensity-score matching.

| **Covariate** | **Before Matching, No. (%)** | | | **After Matching, No. (%)** | | |
|---|---|---|---|---|---|---|
| | Pfizer bivalent cohort (n = 43,216) | Monovalent cohort (n = 100,583) | SMD[a] | Pfizer bivalent cohort (n = 42,327) | Monovalent cohort (n = 42,327) | SMD[a] |
| **Demographics** | | | | | | |
| Age at booster (years)[b] | 73.6 (6.05) | 73.8 (5.90) | 0.04 | 73.6 (6.06) | 73.6 (5.85) | <0.01 |
| Sex | | | | | | |
| Male | 19,726 (45.65) | 45,083 (44.82) | 0.02 | 19,344 (45.70) | 19,436 (45.92) | <0.01 |
| Female | 23,483 (54.34) | 55,499 (55.18) | 0.02 | 22,976 (54.28) | 22,890 (54.08) | <0.01 |
| Race | | | | | | |
| White | 35,382 (81.87) | 73,809 (73.38) | 0.20 | 34,597 (81.74) | 35,104 (82.94) | 0.03 |
| Black or African American | 3,977 (9.20) | 15,559 (15.47) | 0.19 | 3,974 (9.39) | 3,822 (9.03) | 0.01 |
| Asian | 587 (1.36) | 4,786 (4.76) | 0.20 | 587 (1.39) | 426 (1.01) | 0.04 |
| Unknown | 3,074 (7.11) | 5,683 (5.65) | 0.06 | 2,976 (7.03) | 2,800 (6.62) | 0.02 |
| Ethnicity | | | | | | |
| Hispanic or Latino | 3,525 (8.16) | 8,962 (8.91) | 0.03 | 3,422 (8.09) | 2,664 (6.29) | 0.07 |
| Not Hispanic or Latino | 37,827 (87.53) | 87,585 (87.08) | 0.01 | 37,064 (87.57) | 37,506 (88.61) | 0.03 |
| Unknown | 1,864 (4.31) | 4,036 (4.01) | 0.02 | 1,841 (4.35) | 2,157 (5.10) | 0.04 |
| **Diagnoses** | | | | | | |
| Type II diabetes mellitus | 14,862 (34.39) | 23,497 (23.36) | 0.25 | 14,147 (33.42) | 13,775 (32.54) | 0.02 |
| Overweight and obesity | 13,305 (30.79) | 20,119 (20.00) | 0.25 | 12,636 (29.85) | 12,538 (29.62) | <0.01 |
| Disorders of lipoprotein metabolism and other lipidemias | 30,867 (71.43) | 54,559 (54.24) | 0.36 | 29,984 (70.84) | 31,001 (73.24) | 0.05 |
| Mental and behavioral disorders due to psychoactive substance use | 11,553 (26.73) | 12,511 (12.44) | 0.37 | 10,665 (25.20) | 9,660 (22.82) | 0.06 |
| Alcohol related disorders | 2,988 (6.91) | 3,387 (3.37) | 0.16 | 2,848 (6.73) | 2,588 (6.11) | 0.03 |

| | | | | | | |
|---|---|---|---|---|---|---|
| Nicotine dependence | 7,918 (18.32) | 9,319 (9.27) | 0.26 | 7,482 (17.68) | 6,932 (16.38) | 0.03 |
| Essential hypertension | 29,555 (68.39) | 53,712 (53.40) | 0.31 | 28,694 (67.79) | 28,967 (68.44) | 0.01 |
| Ischemic heart disease | 13,104 (30.32) | 22,960 (22.83) | 0.17 | 12,580 (29.72) | 12,446 (29.40) | <0.01 |
| Atrial fibrillation | 5,902 (13.66) | 10,990 (10.93) | 0.08 | 5,720 (13.51) | 5,687 (13.44) | <0.01 |
| Cerebrovascular diseases | 9,605 (22.22) | 15,316 (15.23) | 0.18 | 9,066 (21.42) | 8,862 (20.94) | 0.01 |
| Cerebral infarction | 5,827 (13.48) | 7,277 (7.24) | 0.21 | 5,345 (12.63) | 4,852 (11.46) | 0.04 |
| Persons with potential health hazards related to socioeconomic and psychosocial circumstances | 7,714 (17.85) | 8,077 (8.03) | 0.30 | 6,987 (16.51) | 6,354 (15.01) | 0.04 |
| COVID-19 | 5,959 (13.79) | 4,005 (3.98) | 0.35 | 2,637 (6.46) | 3,710 (9.08) | 0.05 |

[a]Standard mean difference, a metric used to declare imbalance when greater than 0.1.

[b]Average age (standard deviation) is reported for this measurement.

**Survival Analysis**

The Kaplan-Meier Analysis estimates probability of the outcome at a respective time interval (daily time interval is used in this analysis). In order to account for the patients who exited the cohort during the analysis period, and therefore should not be included in the analysis, censoring is applied. In this analysis, patients are removed from the analysis (censored) after the last fact in their record. The proportional hazard assumption was tested using the generalized Schoenfeld approach. The TriNetX Platform calculates the hazard ratios and associated confidence intervals using R's Survival package v3.2-3. For generating hazard ratios, TriNetX sets robust=FALSE using the R survival package, which is a limitation of the TriNetX platform since it does not consider potential clustering of patients within the healthcare organizations or specific geolocations. All Statistical tests were conducted on 1/28/2023 within the TriNetX Analytics Platform with significance set at p-value < 0.05 (two-sided).
